# Detection of iron and zinc in human skin using non-invasive Raman spectrophotometer – A validation study among children under five years of age living in sub-Saharan Africa

**DOI:** 10.64898/2026.04.22.26351546

**Authors:** Carol A. Abidha, Benedicta S. Amevor, Isabel Mank, John Oguso, Michael Mbata, Boubacar Coulibaly, Claudia M. Denkinger, Raissa Sorgho, Ali Sié, Erick M.O. Muok, Ina Danquah

## Abstract

**Background:** Sub-Saharan Africa (SSA) still experiences a high burden of micronutrient deficiencies. For monitoring of micronutrient status among young children in SSA, non-invasive alternatives to blood-based biomarkers are desirable. Handheld Raman spectrophotometry appears to offer this alternative to quantify intracellular stores of micronutrients. In rural Burkina Faso and Kenya, we validated the Cell-/SO-Check device (ZellCheck®) against conventional laboratory-based methods.

**Methods:** For this validation study, we recruited children aged ≥24 months attending routine clinics within the Health and Demographic Surveillance Systems (HDSS) in Siaya and Nouna. Anthropometric measurements and venous blood samples were taken. Plasma ferritin, soluble transferrin receptor (sTfR) and C-reactive protein (CRP) were measured by ELISA, and plasma zinc by atom absorption. The spectrometer was used to quantify zinc and iron. For continuous outcomes, we generated Bland Altman plots and calculated bias and limits of agreement (LoA). For binary outcomes, we produced Receiver Operator Characteristic (ROC) areas under the curve (AUC), and estimated sensitivity, specificity and predictive values.

**Results:** We analysed data of 48 children from Burkina Faso and 54 children from Kenya (male: 53%; age range: 24-66 months). According to spectrophotometry, the proportions of iron deficiency and zinc deficiency were 16.7% and 25.5%, respectively. The median concentrations were for ferritin 24.0 µg/L (range: 2.0-330.0), for sTfR 5.7 mg/L (2.8-51.0), and for zinc 9.9 µmol/L (5.2-25.0). The corresponding bias for iron levels by spectrophotometry was 42.4 with LoA: - 18.7, 103.6. The bias for zinc levels was 7.5 with LoA: −49.3, 64.2. For the classification of deficiency, the ROC-AUC, sensitivity, and specificity for spectrophotometry vs. biomarker-based diagnosis were for iron deficiency 0.62, 68% and 55%, respectively, and for zinc deficiency 0.55, 33% and 91%, respectively.

**Conclusions:** The Cell-/SO-Check device may be used to rank children in population-based studies in SSA according to their zinc status, but not iron status. The method should not replace the standard laboratory measurements for clinical diagnoses of zinc and iron deficiencies.

## Introduction

The global burden of micronutrient deficiencies and undernutrition is still staggering despite the substantial achievement in increasing global food availability [1]. According to the 2022 global nutrition report, all forms of malnutrition are still unacceptably high across the globe [2]. It is estimated that about 2 billion people have one or two micronutrient deficiencies with children being the most affected. Approximately 45% of deaths in children under the age of 5 years is attributed to malnutrition [3]. Although the malnutrition crisis is global, sub-Saharan Africa (SSA) experiences a triple burden, including undernutrition, micronutrient deficiencies, and obesity-related conditions. This has led to an increased prevalence of nutrition-related diseases since nutrition is a key determinant of health and well-being [4].

Climate change is exacerbating this situation, particularly through its direct and indirect impacts on food security [5]. Increased emissions of greenhouse gases reduce the content of iron and zinc in many staple crops in SSA [6,7]. This will likely lead to an increase in micronutrient deficiencies in the most vulnerable population, children below the age of 5 years [6]. In addition, climate change affects the spread of infectious diseases such as malaria in this region, which in turn are linked to micronutrient status [8,9]. Long term, viable interventions that are an integral part of the food systems-approach are necessary to achieve resilience against the impacts of climate change on agriculture and thus improve nutritional status of children.

For regular monitoring of climate-sensitive micronutrients among young children in SSA, non-invasive alternatives to the Gold-standard blood-based biomarkers are desirable. For iron status, the conventional biomarkers are plasma ferritin, soluble transferrin receptor (sTfR), and C-reactive protein (CRP) [10,11]. For zinc status, plasma zinc concentrations by atom absorption or by mass spectrometry are the most common detection methods [12].

Handheld Raman spectrometry appears to be a promising alternative to quantify intracellular stores of climate-sensitive micronutrients, including iron and zinc. This spectrometer measures 20 minerals and micronutrients as well as 14 toxic heavy metals. Its working principle is based on each element reflecting or absorbing light at a certain wavelength. The higher the sample concentration, the more the light is absorbed. The device uses emissions of characteristic frequencies of the specific elements tested [13,14]. However, Raman spectrometry has not been evaluated among young children of SSA origin for the detection of their micronutrient status, and thus, information about the validity of the detection method should be elucidated.

Herein, we validated the Cell-/SO-Check device (ZellCheck®) using conventional laboratory-based methods as the gold standard. We aimed at identifying the potential of using either a handheld Raman spectrometer or laboratory-based methods to measure the status of climate-sensitive micronutrients (iron, zinc) among SSA children aged above 24 months. Specifically, we determined bias and Level of Agreement (LoA) for continuous outcome variables as well as Receiver Operator Characteristic (ROC) Area under the Curve (AuC), sensitivity, specificity and predictive values for dichotomous outcome variables.

## Methods

### Study site and population

The study was conducted within the Health and Demographic Surveillance Systems (HDSS) in Siaya, Kenya and Nouna, Burkina Faso. Participants were recruited between September and October 2020. Children aged ≥24 months living in the areas of the Nouna or the Kisumu HDSS were included. The Nouna HDSS in Northwest Burkina Faso covers rural and semi-urban areas and has a population of about 90,000 individuals. The predominant occupation in this area is subsistence farming [15]. The Kisumu HDSS in Western Kenya has a total of 36 health facilities and monitors a predominantly rural and culturally homogeneous population of about 220,000 individuals. More than 95% of the residents are from the Luo ethnic community; the main economic activities are subsistence farming, fishing and local trading [16].

This study adhered to the principles of the conduct of research laid down in the latest version of the World Medical Association’s Declaration of Helsinki. Particularly, this project adhered to the criteria for including under-aged populations (§20 and §28). Ethical approval was obtained from the responsible ethics committees in Germany, Burkina Faso, and Kenya. Participation in the study was completely voluntary. Written informed consent was provided by caregivers of all study participants. The caregivers were guided through the informed consent form and were provided with all the information about their participation and that of their child(ren). Those who consented were provided with questionnaires asking for the recent medical history of their child(ren). They were also asked to indicate any type of prescribed medicine that their child(ren) was using at that moment.

### Demographic and anthropometric measurements

General demographic information including age (in months) and sex was collected in questionnaire-based interviews. Body weight and length/height measurements of the participating children were taken (SECA213 and SECA874, Germany). Length/height was measured to the nearest cm in duplicates. Within 10 minutes, weight was assessed to the nearest 100 g in duplicates. The mean of the measurements was used in the analysis. Body Mass Index (BMI) was calculated as weight/(height)^2^ in kg/m^2^.

### Blood sample collection, processing, and storage

For each participating child, random venous blood samples were drawn into a 1.5 mL serum tube and into a 1mL heparin tube. Blood groups of the participants were detected on the spot based on Hemagglutination. Plasma and serum were then separated by centrifugation within 3 hours after blood collection and temporarily stored at −20°C. To avoid intra laboratory variation bias, all the collected samples were shipped from Nouna and Kisumu to Heidelberg University Hospital on dry ice, for biomarker analysis. Plasma ferritin, sTfR and CRP concentrations were determined by Enzyme-Linked Immunosorbent Assay (ELISA) kit from Invitrogen, (BMS2004; Carlsbad, CA, USA). Using sTfR, iron deficiency was defined as sTfR >4.4 mg/L for males and >5.0 mg/L for females. Using ferritin and CRP, iron deficiency was defined as ferritin < 15 µg/L if CRP ≤ 5g/dl or ferritin < 30 µg/Lif CRP > 5g/dl. For zinc status, plasma concentrations of zinc were measured by atom absorption. Zinc deficiency was defined as zinc < 7.7µmol/l.

### Measurements using the handheld Raman spectrometer

First, the blood groups (A/B/0, rhesus factor +/-) of the participants were measured using EldonCard according to the manufacturers protocol (EldonCard, Eldon Biologicals A/S, Denmark). Next, participant details including age, sex, weight and height measurements were keyed into the software of the spectrometer before use. The spectrometer was then used on four different points on each participant’s palm. A little pressure was exerted on these four points to send out light using a tungsten diode and an ultraviolet (UV) source, which then detected the corresponding parameters. The resulting measurements are compared with a reference database that contains values from corresponding biopsies.

### Data analysis

The data analysis was conducted using the statistical softwares ***“R”*** (version 3.2.2) and ***MedCalc®*** (version 22.009). A descriptive summary of the study population (n=102) including demographics, anthropometrics, and the micronutrient profiles was conducted using the ***“Table1”*** package. Continuous variables are presented as medians and interquartile ranges (IQR). Categorical data are presented as percentages (n). Further analyses were conducted using only complete cases(n=72), excluding participants with missing data (n=30).

### Analyses of Continuous Outcomes

To assess agreement between spectrometer-based and laboratory-based micronutrient concentrations, Bland–Altman methods were applied. Because spectrometer outputs were expressed as percentages on variable scales including negative values, zinc and ferritin concentrations were rescaled to a 1–100 standard-unit range using the “RPMG” package to facilitate comparison. For each biomarker, we calculated the mean difference (bias) and the 95% limits of agreement (LoA), and generated Bland–Altman plots to visually inspect systematic differences and identify potential outliers. Distributional assumptions for both laboratory and spectrometer measurements were evaluated using the Shapiro–Wilk test. To further characterise the relationship between methods, scatter plots were constructed and Pearson correlation coefficients calculated. Additional exploration of possible proportional bias was conducted using simple linear regression of the paired differences on the mean of the paired measurements.

### Analyses of Binary Outcomes

For binary outcomes representing micronutrient deficiency (iron or zinc), diagnostic accuracy of spectrometer-based classifications was evaluated against biomarker-defined reference standards. Receiver Operating Characteristic (ROC) curves were constructed for each deficiency definition, and the area under the ROC curve (AUC) was estimated to quantify discriminatory performance. Sensitivity, specificity, and positive and negative predictive values were calculated using established biomarker thresholds as the gold standard. Analyses were stratified by deficiency type (iron or zinc) and by the specific laboratory reference measure (ferritin/CRP or sTfR for iron).

## Results

### Characteristics of the study population

A total of 108 children were recruited, of whom 6 were excluded because insufficient blood samples prevented laboratory-based measurement of zinc, ferritin, and sTfR. This resulted in 102 participants with data available for demographic and distributional analyses (**Figure 1**). However, complete biomarker and spectrometer measurements were not available for all participants. Only 72 children had complete paired data for all variables required for the agreement and diagnostic accuracy analyses and therefore constituted the final sample for the deeper method comparison.

**Figure 1:**
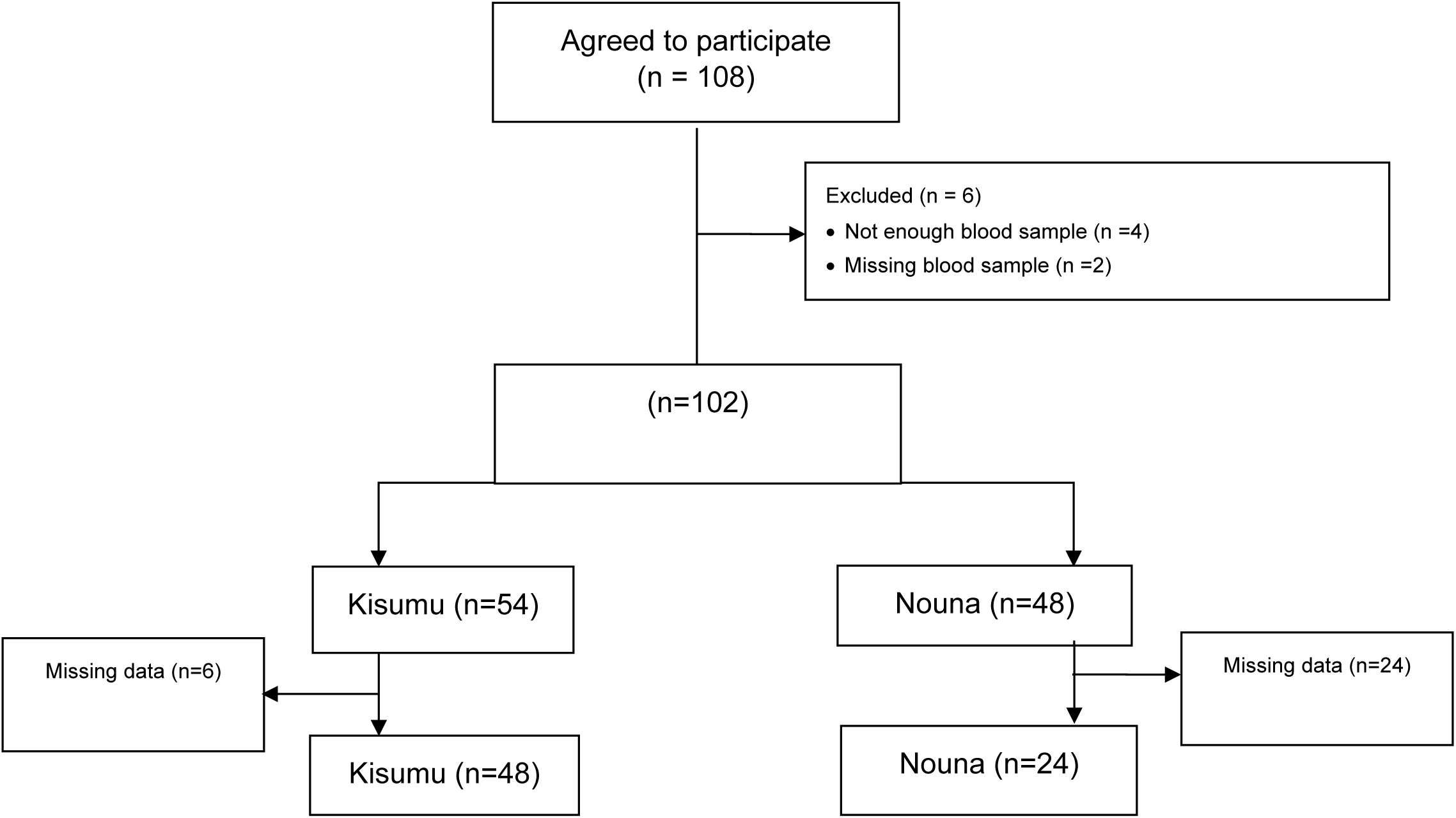
Flow chart of sample selection.

Of the total analytical sample (n=102), 48 came from the Nouna HDSS and 54 children originated from the Kisumu HDSS (**Figure 1**). The sample included 54 males, and 48 females **(Table 1).** The median age of the study participants was 40 months, ranging from 24 to 66 months. The median body weight was 14 kg (range: 10-20 kg), whereas the median height was 96 cm (range: 76-120 cm). Participants in Kenya had higher CRP concentrations (range: 2 mg/l – 110 mg/l) than those in Burkina Faso (range: 2 mg/l– 18 mg/l).

**Table 1.** General characteristics and status of zinc and iron among young children in Kenya and Burkina Faso based on all data set.

|  | **Total** | **Kenya** | **Burkina Faso** |
| --- | --- | --- | --- |
| **N** | 102 | 48 | 54 |
| **Male sex** | 54 (52.9%) | 23 (47.9%) | 31 (57.4%) |
| **Age (months)** | 40 [24, 66] | 43 [24, 63] | 38 [25, 66] |
| **Body weight (kg)** | 14 [10, 20] | 14 [10, 20] | 13 [10, 19] |
| **Length/height (cm)** | 96 [76, 120] | 97 [76, 120] | 95 [79, 120] |
| **Serum zinc (μmol/l)** | 9.9 [5.2, 25] | 9.9 [5.2, 15.0] | 10.0 [6.1, 25.0] |
| **C-reactive protein (g/dl)** | 2.0 [2.0, 110] | 2.8 [2.0, 110] | 2.0 [2.0, 18] |
| **Serum ferritin (μg/l)** | 24 [2.0, 330] | 27 [2.0, 290] | 24 [5.0, 330] |
| **Soluble transferrin receptor (mg/l)** | 5.7 [2.8, 51] | 6.4 [2.8, 51] | 4.6 [2.9, 17] |
| **Zinc deficiency** |  |  |  |
| Zinc < 7.7 μmol/l | 7 (6.9%) | 3 (6.3%) | 4 (7.4%) |
| Missing | 20 (19.6%) | 0 (0%) | 20 (37.0%) |
| Raman spectrophotometry | 26 (25.49%) | 9 (18.75%) | 17 (31.48%) |
| **Iron deficiency** |  |  |  |
| Ferritin < 15 μg/l if CRP ≤ 5 g/dl | 21 (20.6%) | 12 (25.0%) | 9 (16.7%) |
| Ferritin < 30 μg/l if CRP > 5 g/dl | 8 (7.8%) | 6 (12.5%) | 2 (3.7%) |
| sTfR<sub>f</sub> > 4.4 mg/l | 15 (14.7%) | 11 (22.9%) | 4 (7.4%) |
| sTfR<sub>m</sub> > 5.0 mg/l | 15 (14.7%) | 11 (22.9%) | 4 (7.4%) |
| Missing | 43 (42.2%) | 8 (16.7%) | 35 (64.8%) |
| Ferritin + CRP (combined) | 29 (28.4%) | 18 (37.5%) | 11 (20.4%) |
| sTfR (men and women) | 30 (29.4%) | 22 (45.9%) | 8 (14.8%) |
| Raman spectrophotometry | 17 (16.67%) | 7 (14.58%) | 10 (18.52%) |

Data are shown as medians [ranges] and n (percentages).

### Validity of zinc and iron concentrations

The results of iron and zinc status for children according to different measurement methods in the analytical dataset are shown in **Table 2**. For the laboratory-based measurements and the spectrometer-based results, the data were non-normally distributed. Laboratory zinc concentrations ranged between 5.2 and 25.0 µmol/l. This was similar between Burkina Faso and Kenya. Ferritin concentrations ranged between 2.0 and 330.0 µg/l, with no discernible difference between the study sites. Lastly, sTfR concentrations ranged between 2.8 and 51.0 mg/l, showing trends towards higher values in Kenya as compared to Burkina Faso.

**Table 2.**
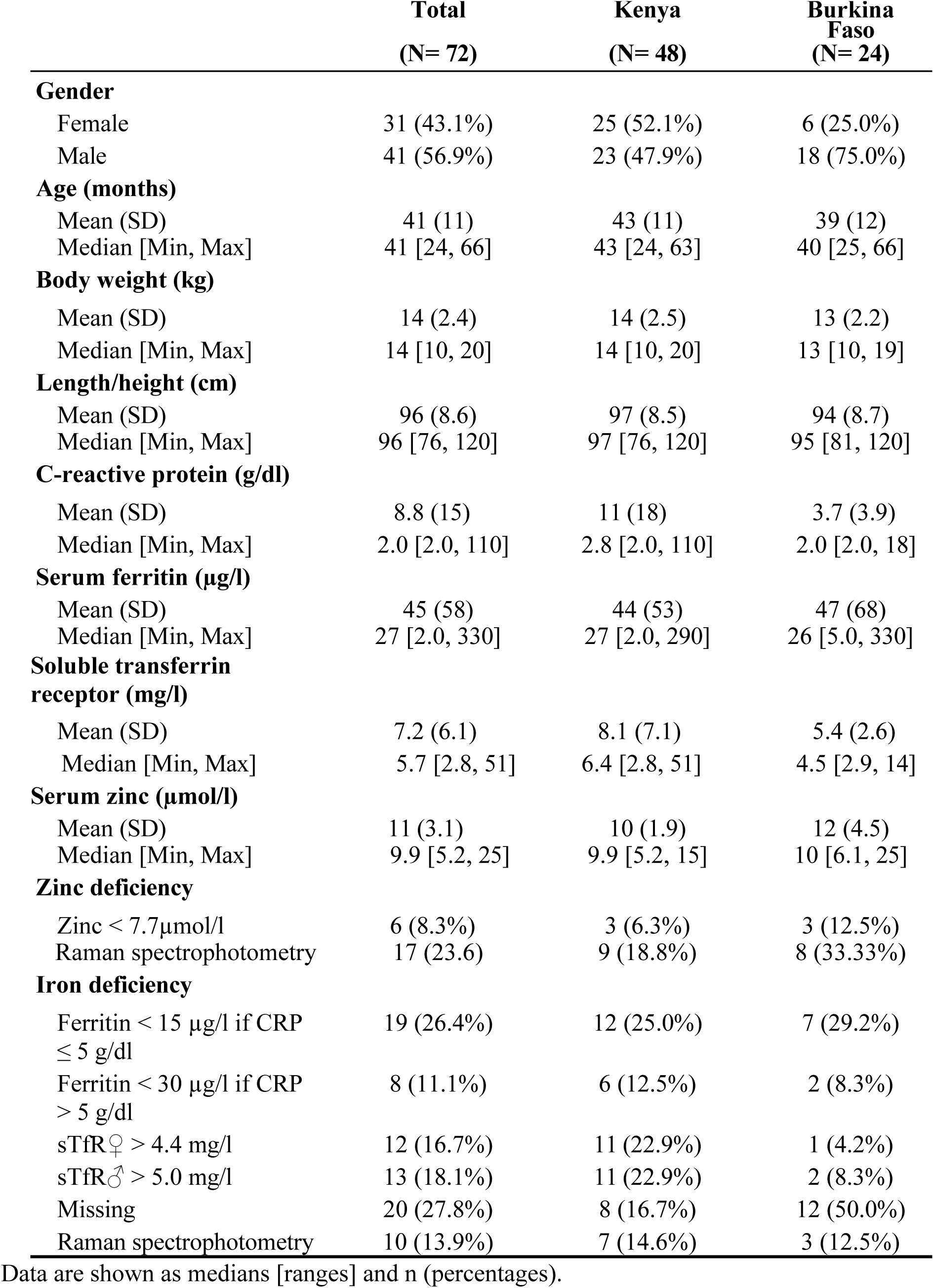
General characteristics and status of zinc and iron among young children in Kenya and Burkina Faso using the complete analysable dataset without missing values (n=72)

**Table 2.** Validity of handheld Raman spectrophotometer for zinc and iron concentrations.

| Comparisons of methods | N | Bias<br>(95% confidence interval) | p-value<br>(Intercept) | 95% Limits of<br>Agreement (LoA) | Accuracy |
| --- | --- | --- | --- | --- | --- |
| Spectrophotometer-based zinc vs. zinc by atom absorption | 72 | 7.48 (0.68, 14.29) | 0.027 | -49.26, 64.23 | ± 56.75 |
| Spectrophotometer-based iron vs. ferritin | 72 | 42.44 (35.11, 49.77) | 0.044 | -18.72, 103.60 | ± 61.16 |
| Spectrophotometer-based iron vs. soluble transferrin receptor (sTfR) | 72 | 46.23 (39.11, 53.35) | 0.738 | -13.19, 105.65 | ± 59.42 |

To compare the different scales and to display the direction, strength, and linearity of the relationships between laboratory-based measurements and spectrometer-based measurements, we re-scaled zinc, ferritin, and sTfR according to the limits of detection from 1 to 100 (standard units) and constructed scatter plots. **Figure 2** presents the scatter plots for spectrometer-based measurements in comparison to laboratory-based measurements. For spectrometer-based zinc vs. atom absorption method (**Figure 2a**), we observed no linear correlation, and the Pearson correlation coefficient was 0.03 (p=0.800). For iron vs. ferritin (**Fig. 2b**), the scatter plot showed no linear correlation, and the Pearson correlation coefficient was 0.01. Also, no linear correlation was observed for iron vs. sTfR (**Figure. 2c**), and the Pearson correlation coefficient was 0.16.

**Figure 2a.**
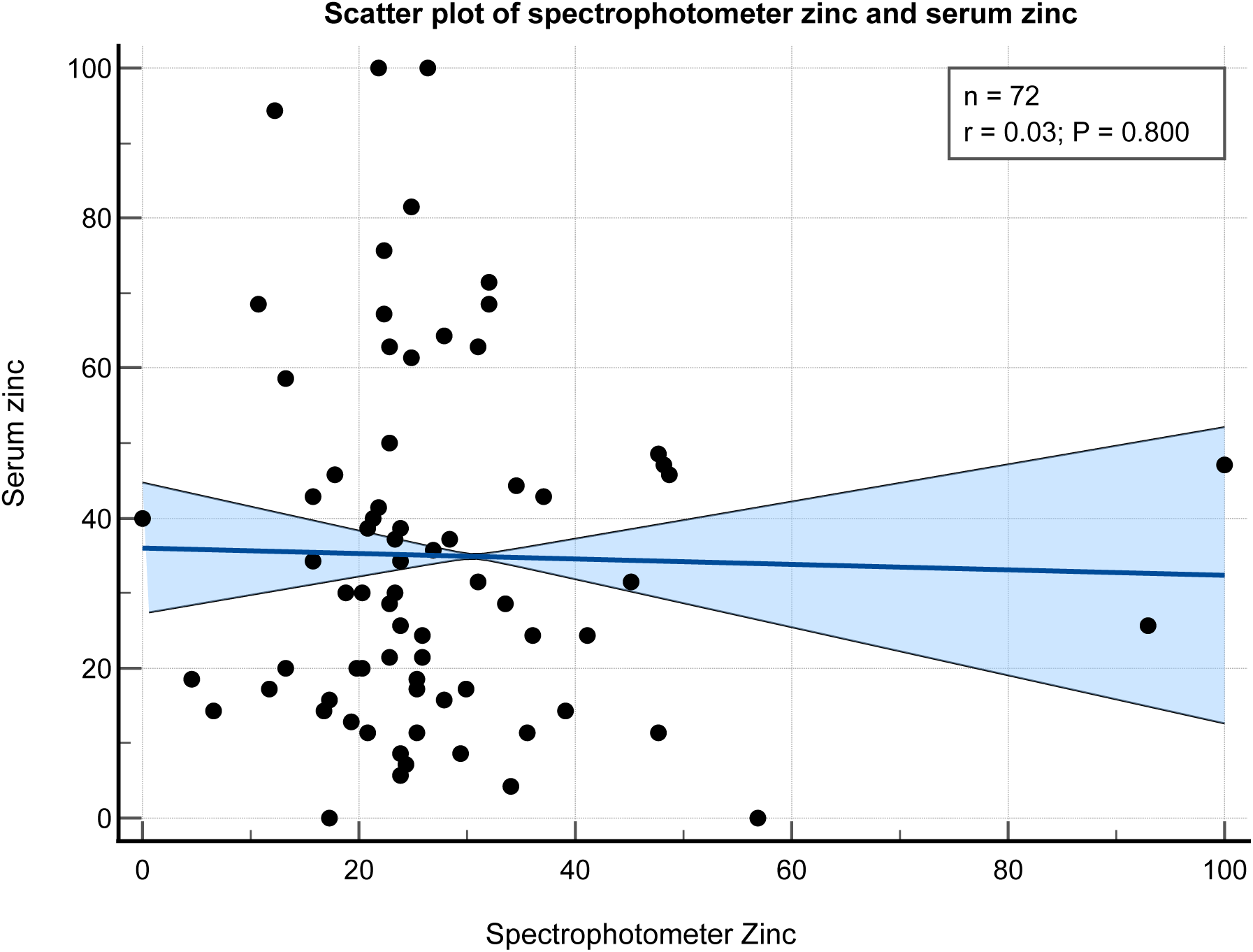
Zinc measurements by handheld Raman spectrometry and laboratory-based measurements. Scatterplot showing spectrophotometer-based zinc measurements (y-axis) plotted against zinc measured by atom absorption (x-axis).

**Figure 2b.**
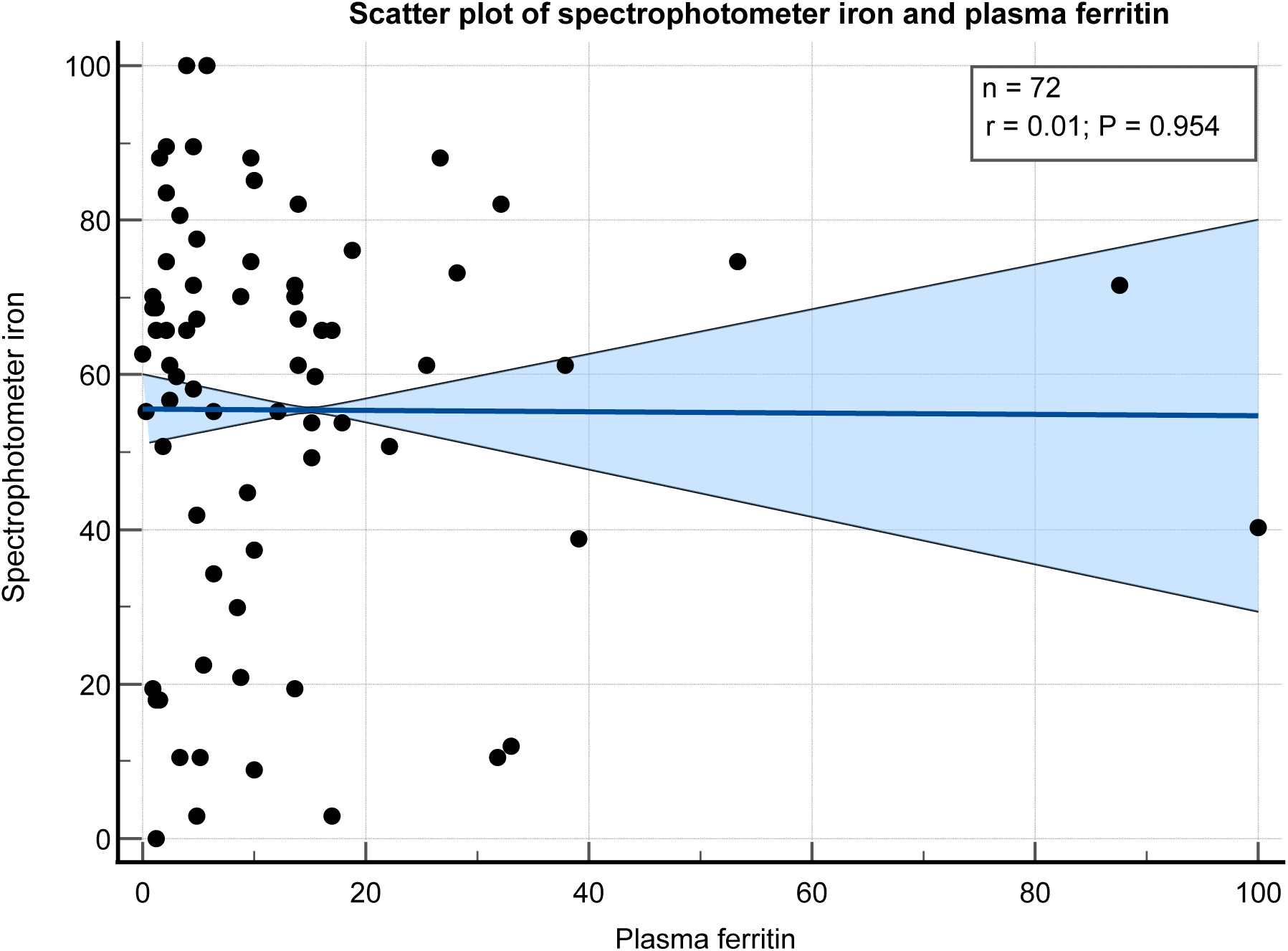
Iron measurements by handheld Raman spectrometry and laboratory-based measurements. (B) Scatterplot showing spectrophotometer-based iron measurements (y-axis) against ferritin measurements (x-axis).

**Figure 2c.**
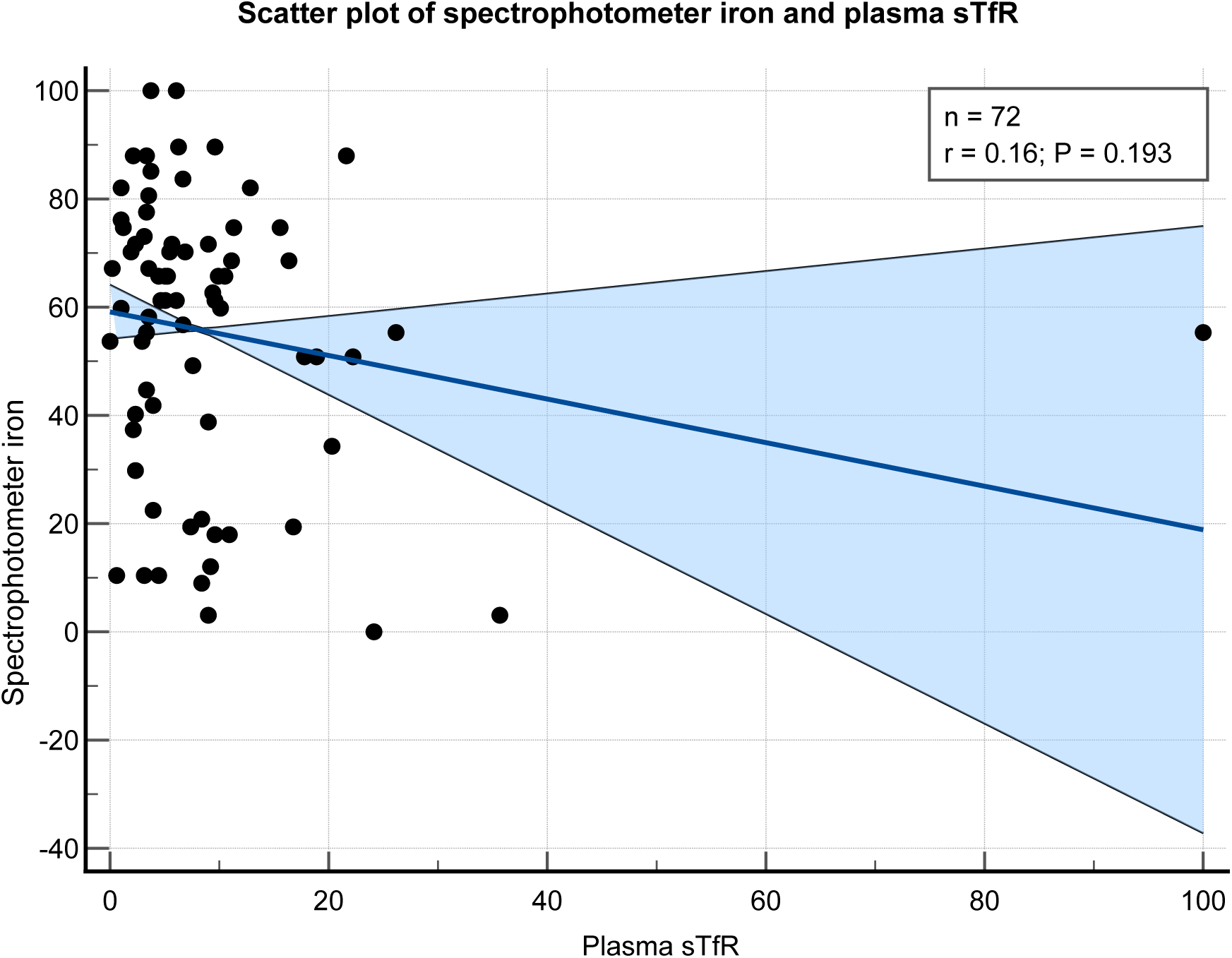
Iron measurements by handheld Raman spectrometry and laboratory-based measurements. Scatterplot showing spectrophotometer-based iron measurements (y-axis) against soluble transferrin receptor (sTfR) measurements (x-axis).

Bland-Altman analysis (**Figure 3**) showed for zinc 95% LoA of −49.26 standard units to 64.23 standard units, and a mean difference (bias) of 7.48 standard units. The corresponding accuracy was 56.75 standard units (**Figure 3a**). For iron vs. ferritin, we observed a 95% LoA of −18.72 standard units to 103.60 standard units, with a mean difference (bias) of 42.44. This translated into an accuracy of 61.16 standard units (**Figure 3b**). For iron vs. sTfR, we observed a 95% LoA of −13.20 standard units to 105.60 standard units, with a mean difference (bias) of 46.20. This translated to an accuracy of 59.40 standard units (**Figure 3c**).

**Figure 3a.**
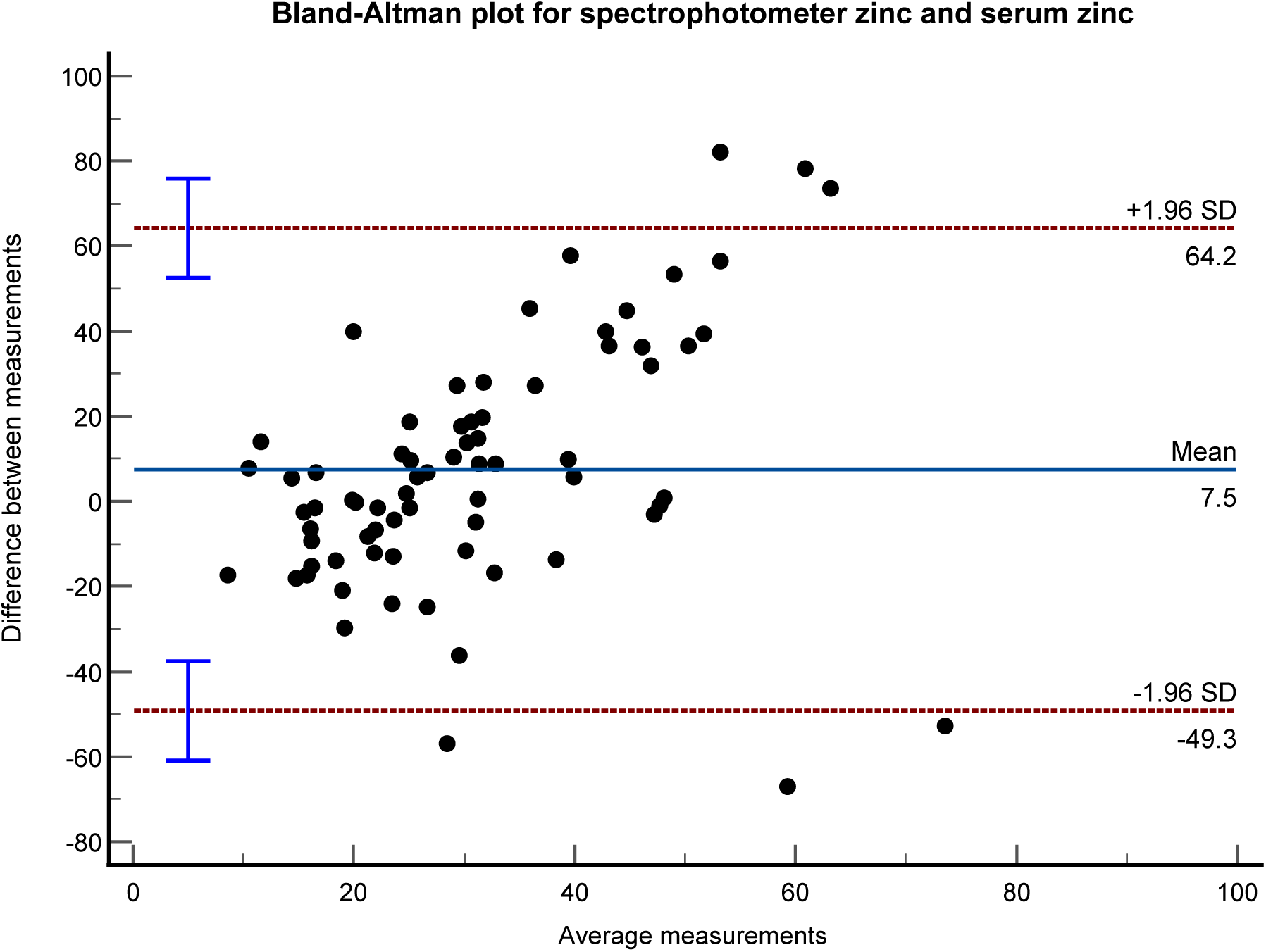
Bland Altman plot depicting agreement between spectrophotometer-based zinc and laboratory zinc concentrations. The blue line represents the mean difference (bias), the red dashed lines represent the upper and lower 95% limits of agreement (LoA), determined as bias ±1.96 standard deviation.

**Figure 3b.**
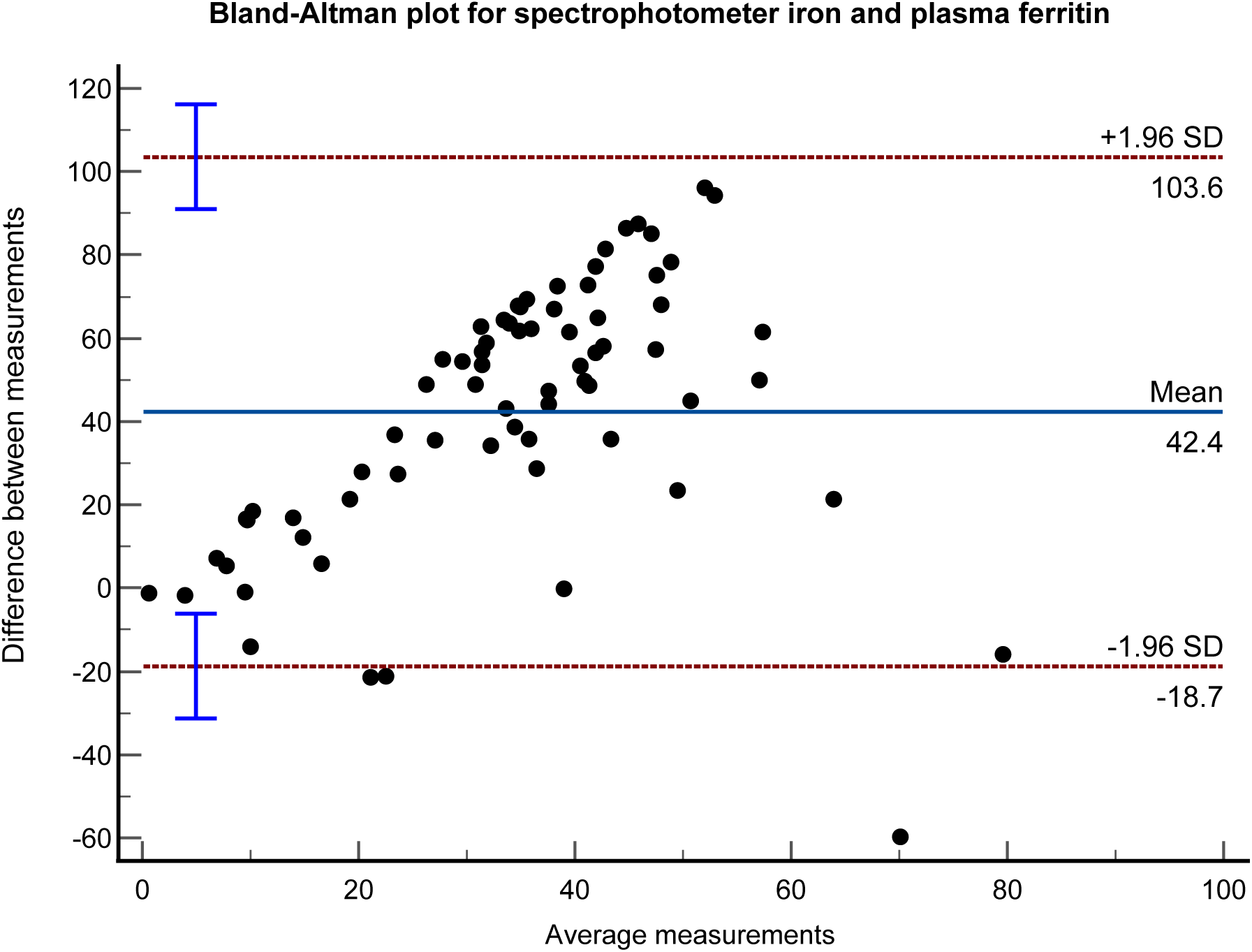
Bland Altman plot depicting agreement between spectrophotometer-based iron and laboratory ferritin concentrations. The blue line represents the mean difference (bias), the red dashed lines represent the upper and lower 95% limits of agreement (LoA), determined as bias ±1.96 standard deviation.

**Figure 3c.**
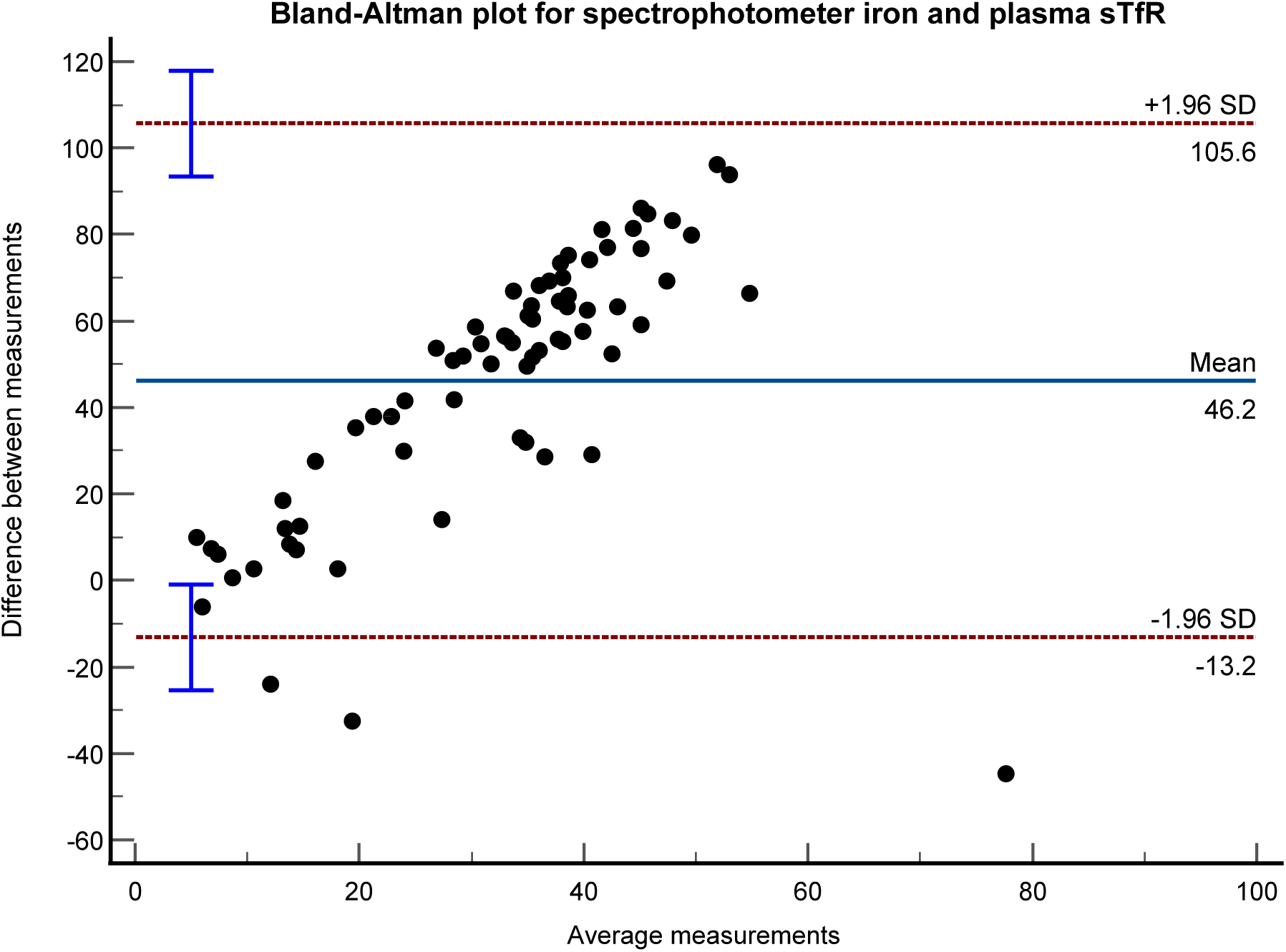
Bland Altman plot depicting agreement between spectrophotometer-based iron and laboratory sTfR concentrations. The blue line represents the mean difference (bias), the red dashed lines represent the upper and lower 95% limits of agreement (LoA), determined as bias ±1.96 standard deviation.

### Validity of zinc and iron deficiencies

We observed the following proportions of children with zinc deficiency and iron deficiency (**Table 2**). When using ferritin and CRP combined, 37.5% of children had iron deficiency; when using sTfR, 34.7% had iron deficiency, and 8.3% had zinc deficiency. The corresponding proportions according to Raman spectrometry were 13.9% for iron deficiency and 23.6% for zinc deficiency.

When using sTfR, the proportions of iron deficiency in Burkina Faso and Kenya were 12.5% and 45.8% respectively. However, when using Ferritin and CRP combined, the proportions were similar between the study sites, i.e 37.5%. The proportions of children with zinc deficiency in Kenya and Burkina Faso were 6.3% and 12.5% respectively. The corresponding proportions of zinc deficiency according to Raman spectrometry were 18.8% and 33.3% respectively.

The results of the ROC-AUC analysis and the validity estimates are presented in **Figure 4** and **Table 3**. We observed moderate predictive ability of the spectrometer-based measurements compared to laboratory diagnosis for zinc and iron, with ROC-AUC ranging from 0.55 for zinc to 0.56 and 0.62 for iron. Further, the specificity of the spectrometer-based analysis was better for zinc (90.1%) than for iron measurements (37.8% and 55.3%). Conversely, the sensitivity was poorer for zinc (33.3%) than for iron measurements (81.5% and 68.0%). These findings translated into poor positive predictive values (PPV) for spectrometer-based measurements compared to biomarkers for both, zinc (25.0%) and iron status (44.0% and 44.7%). At the same time, negative predictive values (NPV) were very good for spectrometer-based zinc measurements (93.8%) but moderate for corresponding iron measurements (77.3% and 76.5%).

**Figure 4a.**
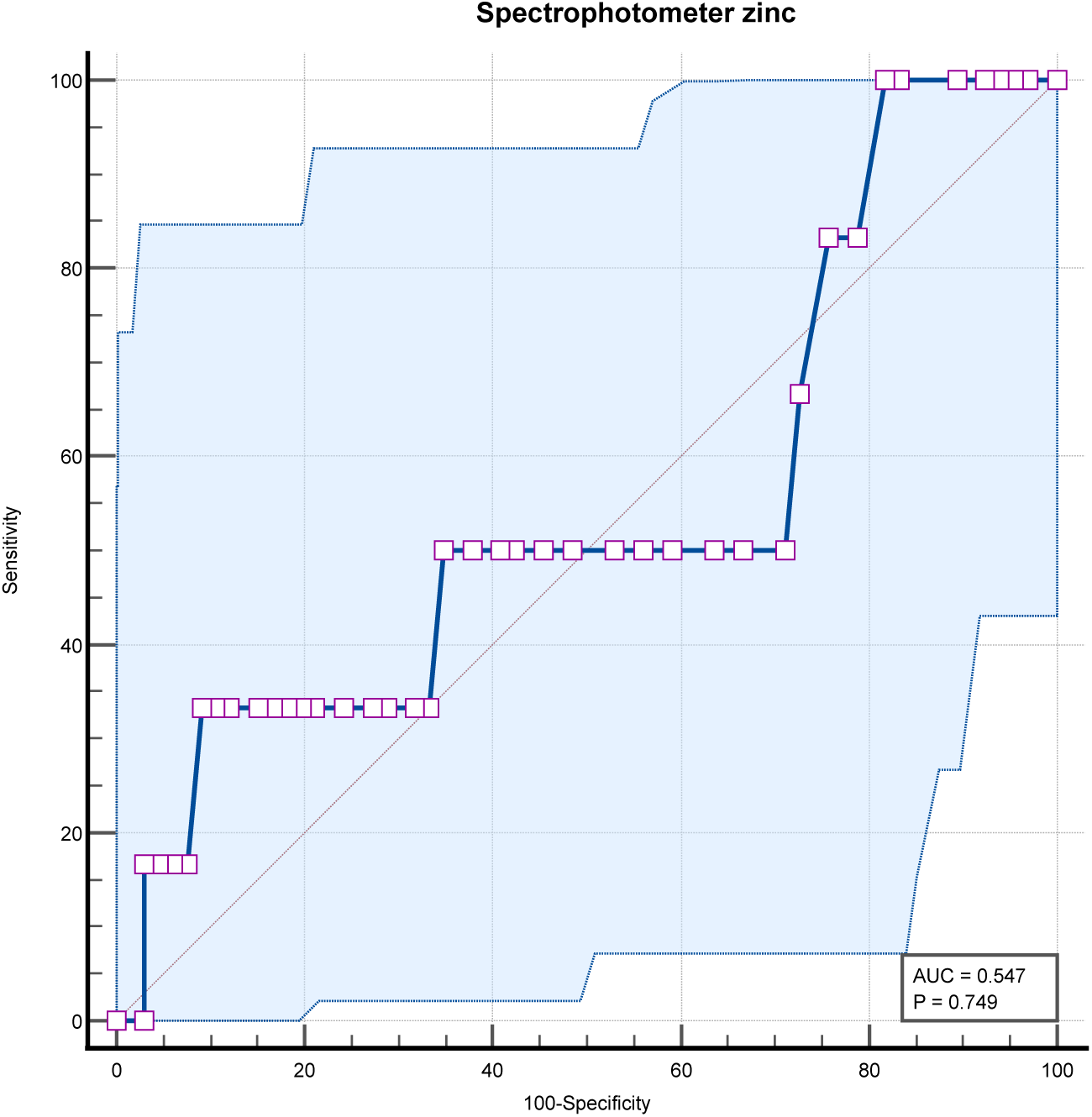
Receiver-operating characteristic (ROC) curve for zinc deficiency. ROC curve for zinc deficiency according to spectrophotometer vs. zinc deficiency according to zincconcentration < 7.7µmol/l.

**Figure 4b.**
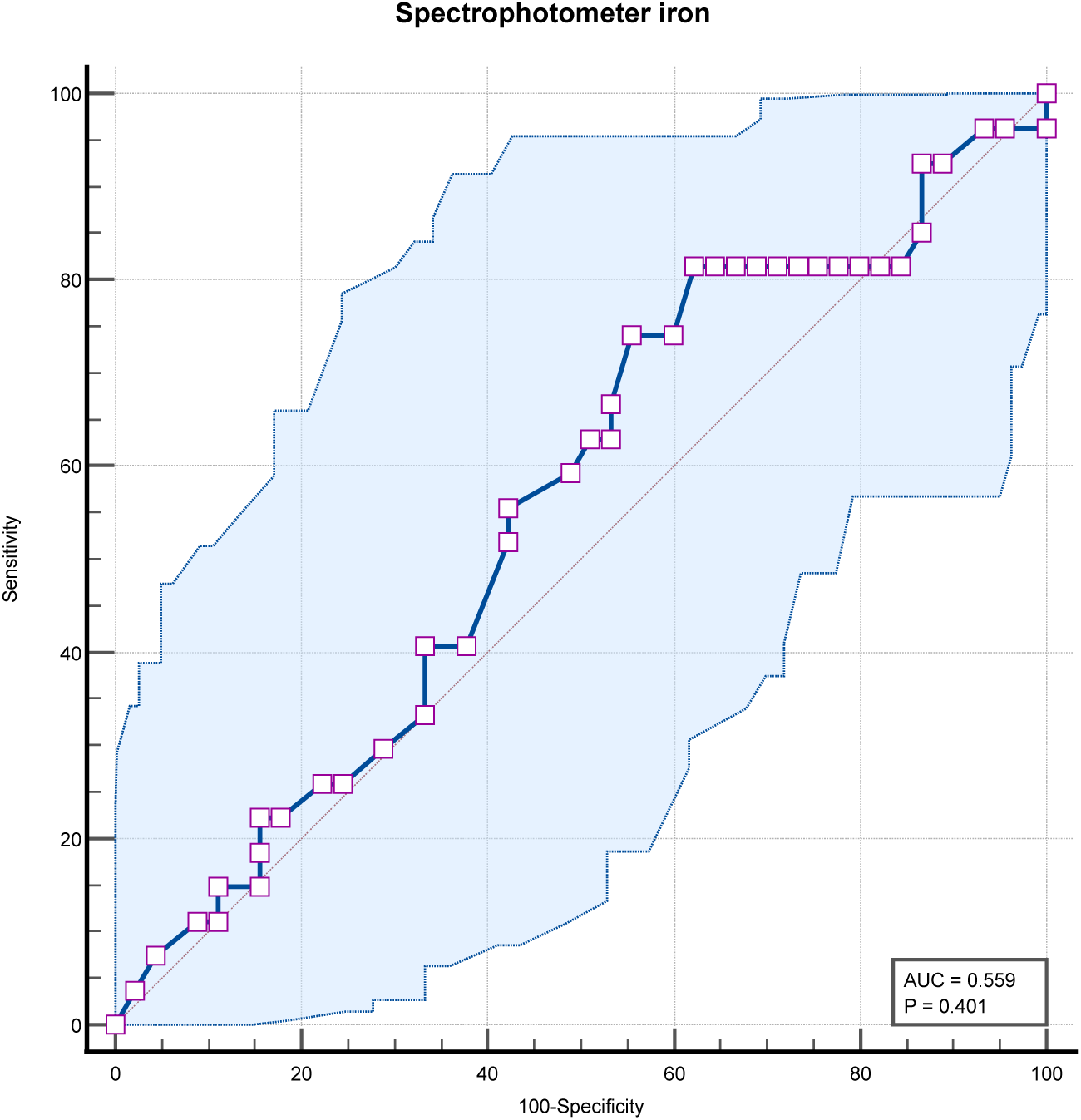
Receiver-operating characteristic (ROC) curve for iron deficiency. ROC curve for iron deficiency according to spectrophotometer vs. iron deficiency according to Ferritin < 15 if CRP ≤ 5g/dl or Ferritin < 30 if CRP > 5 g/dl.

**Figure 4c.**
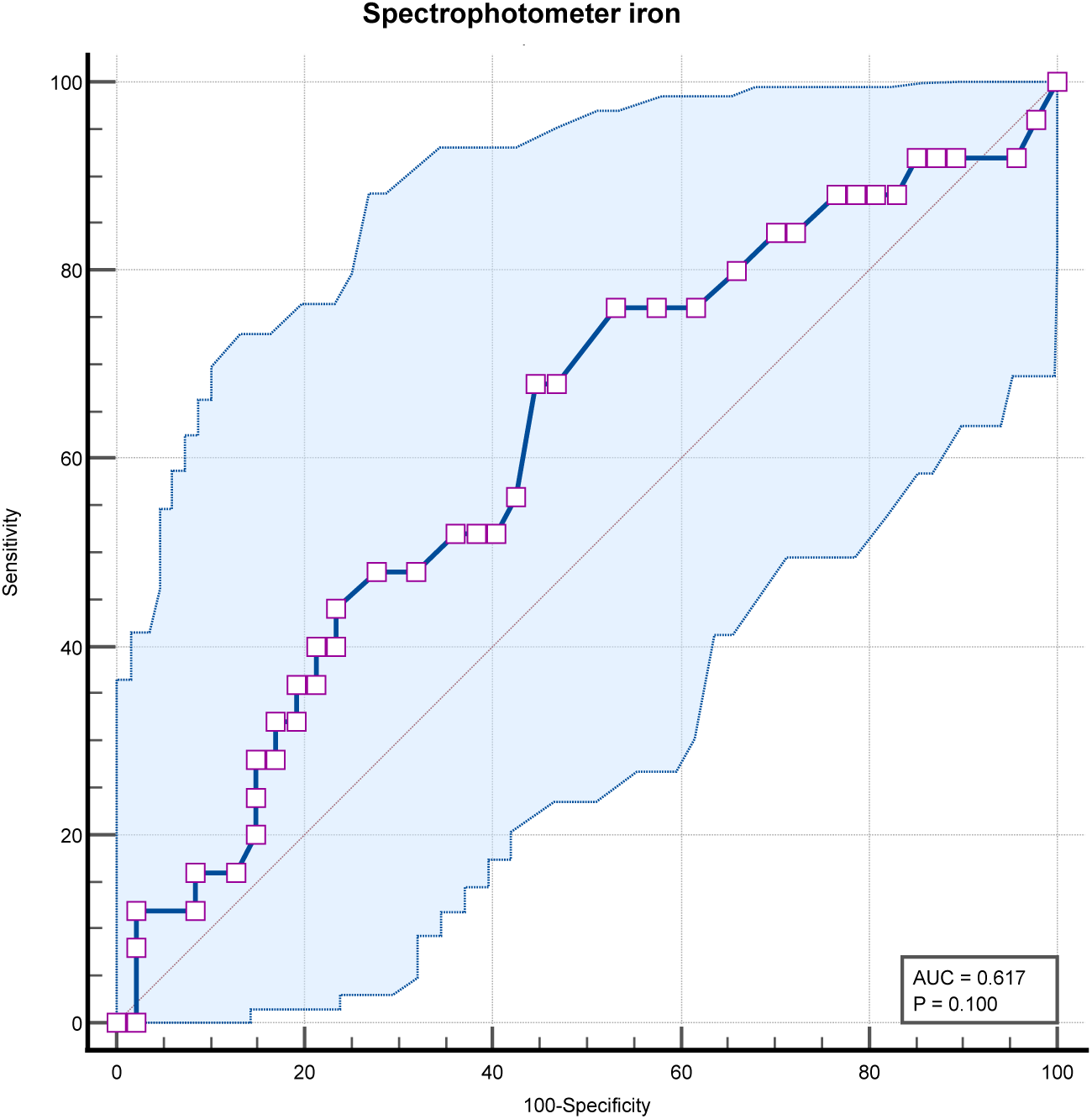
Receiver-operating characteristic (ROC) curve for iron deficiency. ROC curve for iron deficiency according to spectrophotometer vs. iron deficiency according to sTfR♀ > 4.4 mg/l or sTfR♂ > 5.0 mg/l.

**Table 3.** Validity of handheld Raman spectrophotometry for zinc and iron deficiencies.

| Method | Sensitivity (%) | Specificity (%) | PPV (%) | NPV (%) | AUC |
| --- | --- | --- | --- | --- | --- |
| Zinc vs. zinc by atom absorption | 33.33 | 90.91 | 24.99 | 93.75 | 0.547 |
| Iron vs. ferritin | 81.48 | 37.78 | 44.00 | 77.27 | 0.559 |
| Iron vs. sTfR | 68.00 | 55.32 | 44.71 | 76.49 | 0.617 |

## Discussion

### Summary of main findings

This is the first validation study establishing the accuracy of the handheld Raman spectrophotometer Cell-/SO-Check compared to the gold-standard laboratory methods for zinc and iron concentrations. In 102 children under-five from Burkina Faso and Kenya, the continuous measurements of iron showed substantial bias, poor LoA, and low accuracy in comparison to ferritin and sTfR concentrations. These values were better for zinc measurements compared to zinc concentrations by atom absorption. The proportion of zinc deficiency by spectrophotometry was 31.5% in Burkina Faso and 18.8% in Kenya. The proportion of iron deficiency was 18.5% in Burkina Faso and 14.6% in Kenya. The discriminative ability for deficiency by spectrophotometry in comparison to blood-based biomarkers was poor for zinc vs. atom absorption (ROC-AUC: 0.547) and for iron vs. ferritin (ROC-AUC: 0.559); it was moderate for iron vs. sTfR (ROC-AUC: 0.617). This was confirmed by the corresponding results of sensitivity, specificity and predictive values for the test device.

Participants from Kenya had high population-level CRP concentrations, which is consistent with patterns commonly observed in settings with a high burden of infectious diseases. Elevated CRP is frequently seen among children living in environments with intense exposure to infections such as malaria, respiratory tract infections, and diarrhoeal diseases. In such contexts, transient or chronic inflammation is common, and population-level CRP elevations are therefore not unexpected. The higher CRP values observed in our study likely reflect this infectious disease ecology rather than a specific underlying condition, underscoring the importance of accounting for inflammation when interpreting micronutrient biomarkers in similar populations [17,18].

### Zinc and iron deficiencies

Nonetheless, compared to participants from Burkina Faso, almost half of the children in Kenya had iron deficiency based on both Ferritin and CRP levels, and on sTfR. This could explain the elevated CRP levels in participants from Kenya, but is not surprising, because iron deficiency has been proven to be a major public health burden in African children [19]. However, in a study estimating the burden of iron deficiency among African children, iron deficiency among children from Burkina Faso and Kenya was similar [19]. Even though zinc deficiency was low in both settings in the present study, participants from Burkina Faso had a relatively higher serum zinc levels compared to those from Kenya. Nutritionally, this bears the question of what is being done right in Burkina Faso that Kenya can learn from? At the community level, there has been an acknowledged progress and success in the coverage and dispensation of quality nutrition specific and nutrition sensitive programs in Burkina Faso, which likely explains the observed improvement of nutrient profile of Burkinabe children [20]. Still, iron deficiency has a detrimental effect on the health of children and the complex interactions of its biomarkers with inflammation and infections such as malaria should be examined to estimate its true burden in Kisumu [19,21].

### Accuracy of handheld Raman spectrophotometry

The average discrepancy between the spectrometer and laboratory measurements of ferritin was large, indicating an existence of bias. On average, the Spectrometer tests ferritin to be 42.44 times higher than the laboratory measurements with 95% of the differences in measurements between the two methods falling in the range of −18.72 and 103.60. This large discrepancy can lead to false positives for ferritin overload. For zinc, on average, the Spectrometer tests zinc to be 7.48 times higher than the laboratory measurements and 95% of the differences in measurements between the two methods fall in the range of −49.26 and 64.23, which also indicates bias. The range is wide making the results ambiguous. If the limits were narrow (and the bias small), then the two methods would be essentially equivalent. Moreover, The AUC values at 95% confidence interval ranged from 0.55 to 0.62 for spectrometer-based zinc/iron vs. zinc by atom absorption and ferritin, indicating that the spectrometer tests are no better than chance at distinguishing between diseased and non-diseased individuals. This means the spectrometer tests are not clinically adequate [22].

Nevertheless, the P-value for the paired samples t-test of both zinc and ferritin was significant (>0.05), suggesting that the bias albeit present, was consistent. This does not necessarily stipulate that the spectrometer and laboratory methods are incomparable. It could be a matter of calibration discrepancies. In fact, according to Bland & Altman, consistent bias can be adjusted for by subtracting the mean difference of the ferritin or zinc measurements by the spectrometer and the laboratory method from the measurement by the spectrometer method [23].

In addition, for zinc, the spectrometer test correctly identified 90.91% of individuals who did not have zinc deficiency. This is particularly useful for ruling in a diagnosis, as a positive result is more likely to be accurate. The negative predictive value for zinc further confirmed that 93% of individuals who tested negative for a zinc deficiency using the Raman spectrometer are truly not zinc deficient, suggesting that the test is quite accurate in ruling out zinc deficiency [24]. For iron, the spectrometer correctly identified 81.48% of individuals who had iron deficiency as positive. This is desirable because it minimizes the number of false negatives, ensuring that fewer individuals with iron deficiency are missed. A good sensitivity is desirable, but not at the expense of specificity [25].

### Implications for research and public health

Further, the handheld Raman spectrometer uses emissions of characteristic frequencies of the specific compounds tested, a technique that has been widely verified for material analyses [13,14,26]. For biomarker analyses, this method has been well-established and previously validated for detection of both human and animal carotenoid status [27–30]. In fact, the Raman scattering method has been verified to reflect the presence of carotenoids in human skin and to be highly reproducible [30]. These findings highlight the potential of the handheld Raman spectrometer in detecting micronutrient levels. However, the readings obtained from the spectrophotometer have no units of measurement for micronutrients, minerals, and heavy metals. Instead, the read-outs are relative values in reference to biopsy samples. When the investigator has no access to this reference database, it is impossible to truly quantify tissue concentrations of the elements of interest – a major limitation of the device.

For diagnostic and screening purposes, in the clinical setting to guide public health interventions, accurate determination of iron and zinc status is crucial. For the determination of iron deficiency and iron overload based on ferritin levels, there are pre-established cut offs for healthy individuals and for individuals with infection or inflammation [10,31]. It is therefore important that diagnostic tests are accurate enough to detect both zinc and iron deficiency or overload, to initiate treatment. For epidemiological studies determining prevalence, distribution of iron or zinc deficiency and determination of risk of overload in a population, the cut-offs are less stringent [10], and the Raman spectrometry can, with adjustments be utilized [32], as it is time saving, non-invasive and patient friendly.

## Conclusion

This is the first study validating the Raman spectrophotometer Cell-/SO-Check for zinc and iron detection in two SSA populations. While the non-invasive handheld spectrophotometer can be safely used, it should not replace the standard laboratory measurements for clinical diagnoses of zinc and iron deficiencies. The biomarker-based measurements in Burkina Faso and Kenya confirm that considerable proportions of children have iron and zinc deficiency. Thus, more effective public health interventions are needed in SSA to improve iron and zinc status of young children. In addition, underlying common infections in children create difficulties in interpreting biomarkers for iron deficiency. This calls for investments into novel assessment methods that are not confounded by inflammatory state.

## Data Availability

The datasets generated and/or analysed during the current study are available from the corresponding author on reasonable request.

## Acknowledgements

We thank the clinical, field, and lab staff at Agogo Presbyterian Hospital for their assistance with sample collection and processing during the study period. We thank the participants who committed their valuable time and provided biological samples. This study is published with permission of the Director of HIGH.

